# Early Detection of Erythropoietic Protoporphyria Using Sequential Machine Learning on Longitudinal Electronic Health Records

**DOI:** 10.64898/2026.08.15.26360514

**Authors:** Aryan Ayati, Goktug Onal, Alana Sur, Shadera Azzam, Bruce Wang, Vivek A. Rudrapatna

## Abstract

**Objective:** Erythropoietic protoporphyria (EPP) is a rare photodermatosis marked by multi-year diagnostic delays. We developed and externally validated machine learning models to identify patients with EPP earlier from longitudinal electronic health record (EHR) data and estimate undiagnosed disease burden.

**Materials and Methods:** In a retrospective case–control study at two San Francisco health systems—an academic referral center (UCSF) and a safety-net hospital (ZSFG)—we identified 74 confirmed EPP cases using combined diagnostic coding, biochemical criteria, and specialty chart review. Symptom-enriched controls were sampled at a 40:1 ratio. Longitudinal diagnoses, laboratory results, medications, procedures, and encounters preceding the outcome date were modeled with a gradient-boosting classifier (CatBoost) and a state-space sequence model (MAMBA). The best model was deployed across the UCSF population and externally validated at ZSFG without retraining.

**Results:** On the UCSF held-out test set (n=1,865; 43 cases), MAMBA outperformed CatBoost (AUC–ROC 0.91 vs 0.89; average precision 0.42 vs 0.27; precision 65% vs 20%), flagging cases a median of 229 days before documented diagnosis. Deployed across 297,967 symptom-compatible patients, it identified 310 high-risk individuals, implying a prevalence approaching genetic estimates. External validation at ZSFG showed attenuated performance (AUC–ROC 0.72; average precision 0.10) while preserving early detection (median 264 days).

**Discussion:** A sequence model integrating temporal EHR signals detected EPP months before clinical recognition, corroborating genetic evidence of substantial underdiagnosis. Cross-site attenuation reflects population and documentation differences and underscores the need for local recalibration.

**Conclusion:** Longitudinal EHR-based machine learning can shorten EPP diagnostic delay and prioritize patients for confirmatory testing, supporting proactive rare-disease case finding.

## 1. Introduction

Erythropoietic protoporphyria and X-linked protoporphyria are rare inherited disorders of heme biosynthesis characterized by severe non-blistering cutaneous photosensitivity. Erythropoietic protoporphyria is caused by partial deficiency of ferrochelatase (FECH), while X-linked protopophyria arises from gain-of-function mutations in *ALAS2*, both of which lead to abnormal accumulation of protoporphyrin IX in erythrocytes, plasma, and peripheral tissues.^1–3^ Clinically, the two conditions (hereafter referred to as EPP) present with identical clinical symptoms that typically begins in early childhood, with burning pain, erythema, and edema occurring within minutes of sunlight or visible light exposure. A subset of patients develops progressive hepatobiliary complications, including cholestasis, cholelithiasis, and in rare cases, protoporphyric liver failure requiring transplantation.^2–4^

Despite a characteristic symptom profile, EPP is notoriously difficult to diagnose. Cutaneous symptoms occur without consistent visible skin findings, and early presentations often overlap with unrelated conditions such as atopic dermatitis, urticaria, functional abdominal pain, and iron deficiency anemia.^2–4^ Published cohorts consistently describe multi-year or even multi-decade diagnostic delays, during which patients cycle through primary care, dermatology, gastroenterology, and hematology services.^5–6^ These delays carry clinical consequences: avoidable pain, psychosocial burden, unnecessary testing, and progression of hepatic disease that might have been identified with earlier surveillance.

Converging lines of evidence indicate that the rarity of EPP has been overestimated by traditional clinical case counting. A population-scale analysis using the UK Biobank estimated a genetic prevalence of FECH mutations of approximately 1 in 17,000—roughly an order of magnitude higher than clinical estimates of 1 in 75,000 to 1 in 200,000—suggesting that many individuals with pathogenic FECH variants remain undiagnosed in routine care.^7^ This gap between genetic and clinical prevalence highlights an opportunity for systematic case finding using real-world clinical data.

Modern longitudinal electronic health records (EHRs) are well suited to this task. They capture time-stamped diagnoses, laboratory values, procedures, medications, and encounter patterns that, in aggregate, trace a patient’s clinical trajectory over years. Machine learning approaches applied to similar data have successfully identified phenotypic signatures of other rare diseases, including acute hepatic porphyrias, primary immunodeficiencies, and inborn errors of metabolism, typically months before formal diagnosis.^8–11^ However, published work specific to EPP remains limited, and few studies have evaluated generalization across health systems that serve distinct patient populations.

In this study, we developed and externally validated machine learning models to identify patients with EPP using longitudinal EHR data from two San Francisco hospitals: a tertiary academic referral center (UCSF Medical Center) and a public safety-net hospital (Zuckerberg San Francisco General Hospital, ZSFG). Our objectives were to (1) develop predictive models that can identify EPP earlier in the patient’s trajectory; (2) deploy the best-performing model across the UCSF general population to estimate the scale of undiagnosed disease; and (3) externally validate model performance in a demographically distinct safety-net setting.

## 2. Methods

### 2.1 Study Design and Setting

We utilized a retrospective case–control design using longitudinal EHR data from two urban health systems in San Francisco, California. UCSF Medical Center is a tertiary referral academic center with a diverse insured and commercially referred population, and it serves as a regional hub for rare disease expertise, including porphyria evaluation and management. ZSFG is an affiliated public safety-net hospital providing care predominantly to low-income, uninsured, and publicly insured patients with a high prevalence of adverse social determinants of health. The study window included all patients with at least one clinical encounter within the preceding three years at each institution. The study was reviewed by the UCSF Institutional Review Board and determined to be minimal risk; waiver of informed consent was granted because all data were de-identified prior to analysis.

### 2.2 Data Sources

Data were extracted from institutional de-identified clinical data warehouses containing structured EHR elements recorded during routine clinical care. Extracted domains included demographic characteristics, ICD-9 and ICD-10 diagnosis codes, laboratory results with units and timestamps, medication orders, procedures, imaging studies, and encounter-level metadata. For laboratory values relevant to EPP diagnosis, erythrocyte protoporphyrin and plasma protoporphyrin measurements were pulled from the laboratory information system and linked to patient records.

### 2.3 Case Ascertainment

Patients with EPP were identified at UCSF using a comprehensive strategy intended to mitigate known limitations of administrative coding. First, individuals with ICD-9 or ICD-10 diagnosis codes specific to EPP or related porphyrias were identified. Second, because administrative coding is often incomplete or delayed, we incorporated EPP-specific biochemical criteria: patients with documented elevated erythrocyte protoporphyrin levels or protoporphyrin fractionation patterns consistent with EPP were included regardless of ICD status. Third, when diagnostic ambiguity remained, specialty clinic records in dermatology, hematology, and hepatology were reviewed to confirm the diagnosis.

Using this strategy, 74 patients with confirmed EPP were identified. The index date for each case was defined as the earliest documentation of confirmed EPP, based on biochemical testing or formal clinical diagnosis, whichever occurred first.

### 2.4 Control Selection

Controls were drawn from the general UCSF population and were required to have (a) no documented EPP diagnosis on any ICD code and (b) no confirmatory EPP laboratory finding. To construct a clinically meaningful comparison group, controls were further required to carry at least one diagnosis associated with a symptom potentially compatible with EPP, including abdominal pain, iron deficiency anemia or other anemias, rash, pruritus, visual disturbances, liver disease, cholelithiasis and other gallbladder disease, urticaria, or other photosensitivity-related dermatologic conditions (Table 1). This design is intended to simulate the clinically relevant test set on which a deployed model would be expected to discriminate, rather than an artificially easy population of healthy individuals (Figure 2).

**Table 1.**
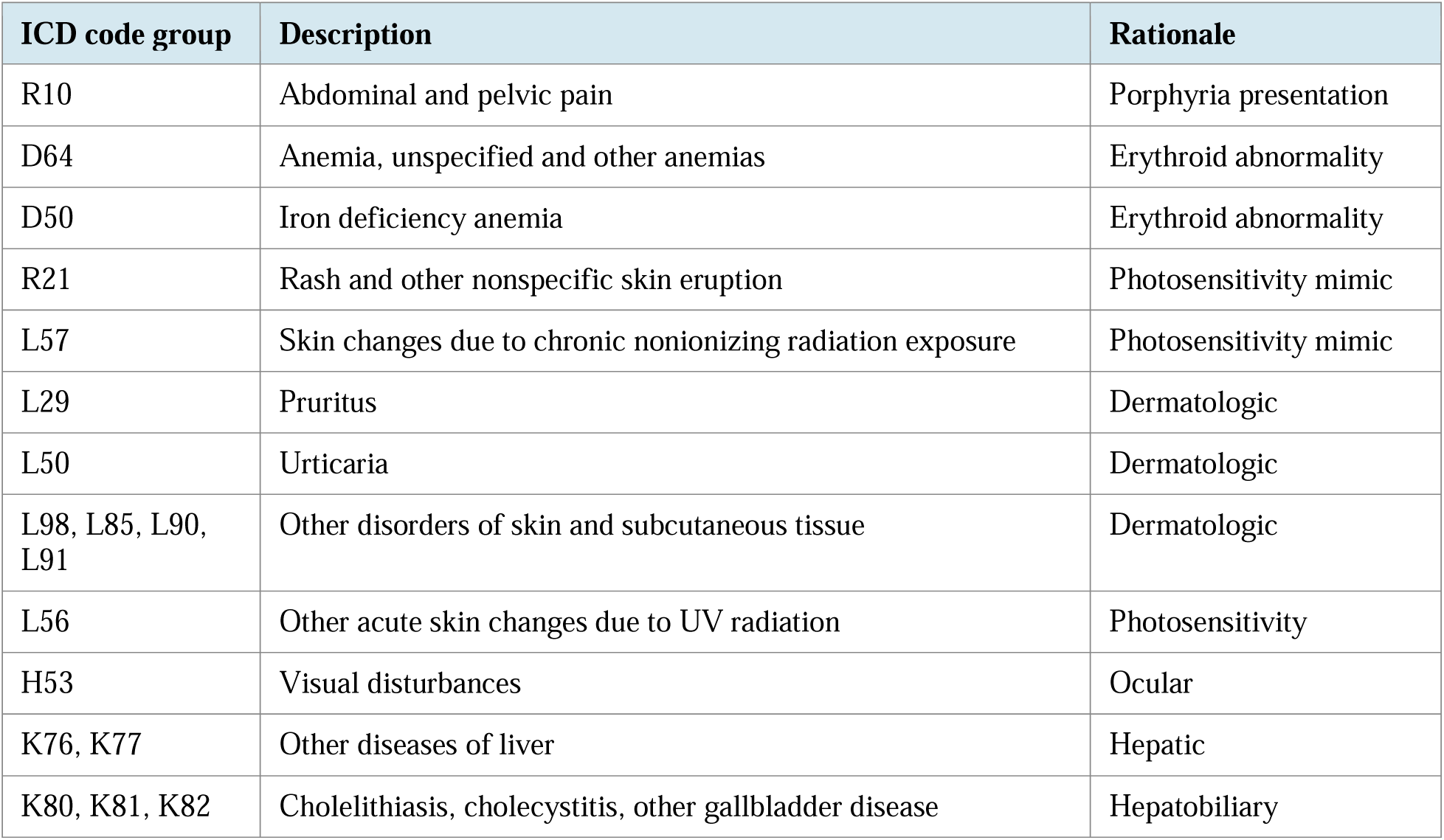
ICD code groups used to define EPP-compatible symptoms for control eligibility and general-population deployment.

| ICD code group | Description | Rationale |
| --- | --- | --- |
| R10 | Abdominal and pelvic pain | Porphyria presentation |
| D64 | Anemia, unspecified and other anemias | Erythroid abnormality |
| D50 | Iron deficiency anemia | Erythroid abnormality |
| R21 | Rash and other nonspecific skin eruption | Photosensitivity mimic |
| L57 | Skin changes due to chronic nonionizing radiation exposure | Photosensitivity mimic |
| L29 | Pruritus | Dermatologic |
| L50 | Urticaria | Dermatologic |
| L98, L85, L90, L91 | Other disorders of skin and subcutaneous tissue | Dermatologic |
| L56 | Other acute skin changes due to UV radiation | Photosensitivity |
| H53 | Visual disturbances | Ocular |
| K76, K77 | Other diseases of liver | Hepatic |
| K80, K81, K82 | Cholelithiasis, cholecystitis, other gallbladder disease | Hepatobiliary |

A 40:1 control-to-case ratio was used for model development to approximate the relative imbalance expected at deployment while preserving model training stability. For controls, the index date was defined as the date of last clinical encounter within the study window.

### 2.5 Feature Extraction and Variable Construction

For each patient, longitudinal EHR data were extracted from the first recorded encounter with potential EPP symptoms until the outcome date. The outcome date was defined as the date of confirmatory EPP diagnosis for cases, and the date of last recorded encounter (or death, where applicable) for controls. Given the rarity of EPP, controls with no EPP detected as of their last encounter were assumed to be true non-cases. All features were restricted to information available strictly prior to the outcome date, ensuring temporal validity and preventing leakage of diagnostic information into the predictor set. Feature domains included demographic characteristics (age, sex, race/ethnicity), all ICD-9 and ICD-10 diagnoses, laboratory results, medications, procedures, imaging studies, and encounter types.

To capture temporal dynamics, running cumulative counts of diagnosis codes and cumulative laboratory summaries were generated across the patient’s longitudinal record. These longitudinal features were recalculated at every historical encounter, so that the resulting dataset was structured in long format: each patient-encounter timestamp corresponded to a single row, with feature values reflecting only the information available up to that point in time. Each row, therefore, represented a distinct opportunity for the model to synthesize the accumulated clinical history and update its prediction of future EPP status, rather than yielding one static prediction per patient. Feature selection was performed using L1-penalized (Lasso) logistic regression to reduce dimensionality and retain informative predictors.

### 2.6 Model Development

The UCSF analytic cohort was divided into training (80%), validation (10%), and independent test (10%) sets using stratified sampling to preserve class balance across splits. Two modeling approaches were evaluated in parallel: (1) CatBoost, a gradient boosting decision tree algorithm with native handling of categorical variables and strong performance on structured clinical tabular data;_12_ and (2) MAMBA, a selective state-space sequence architecture designed for efficient modeling of long-range temporal dependencies in sequential data.^13^ The MAMBA model was trained on patient-level event sequences that preserved the temporal order of each clinical event. Five-fold cross-validation was performed within the training set for both models, and hyperparameters were tuned based on validation-set performance.

### 2.7 Performance Evaluation

Model performance on the held-out test set was evaluated using accuracy, sensitivity (recall), specificity, precision (positive predictive value), F1 score, area under the receiver operating characteristic curve (AUC–ROC), and area under the precision–recall curve (average precision, AP). Given the rarity of EPP and the resulting class imbalance, precision–recall performance was prioritized over overall accuracy when comparing models. Operating thresholds for binarized predictions were selected on the validation set and then applied, without modification, to the test set.

To estimate potential clinical impact, we computed the median time difference between the model’s first high-risk prediction and the documented diagnosis date among true-positive cases. This quantity, which we refer to as diagnostic lead time, estimates how much earlier a case could theoretically have been flagged by the model under routine deployment.

### 2.8 Population-Level Deployment

The best-performing model was then applied, without retraining, to the broader UCSF population. To match the symptom-enriched development cohort, deployment was restricted to patients with any encounter during the three-year study window who carried at least one EPP-compatible symptom diagnosis (N = 297,967). The flagged subset was interpreted as a prioritized candidate list for prospective confirmatory erythrocyte protoporphyrin testing (Figure 1).

**Figure 1.**
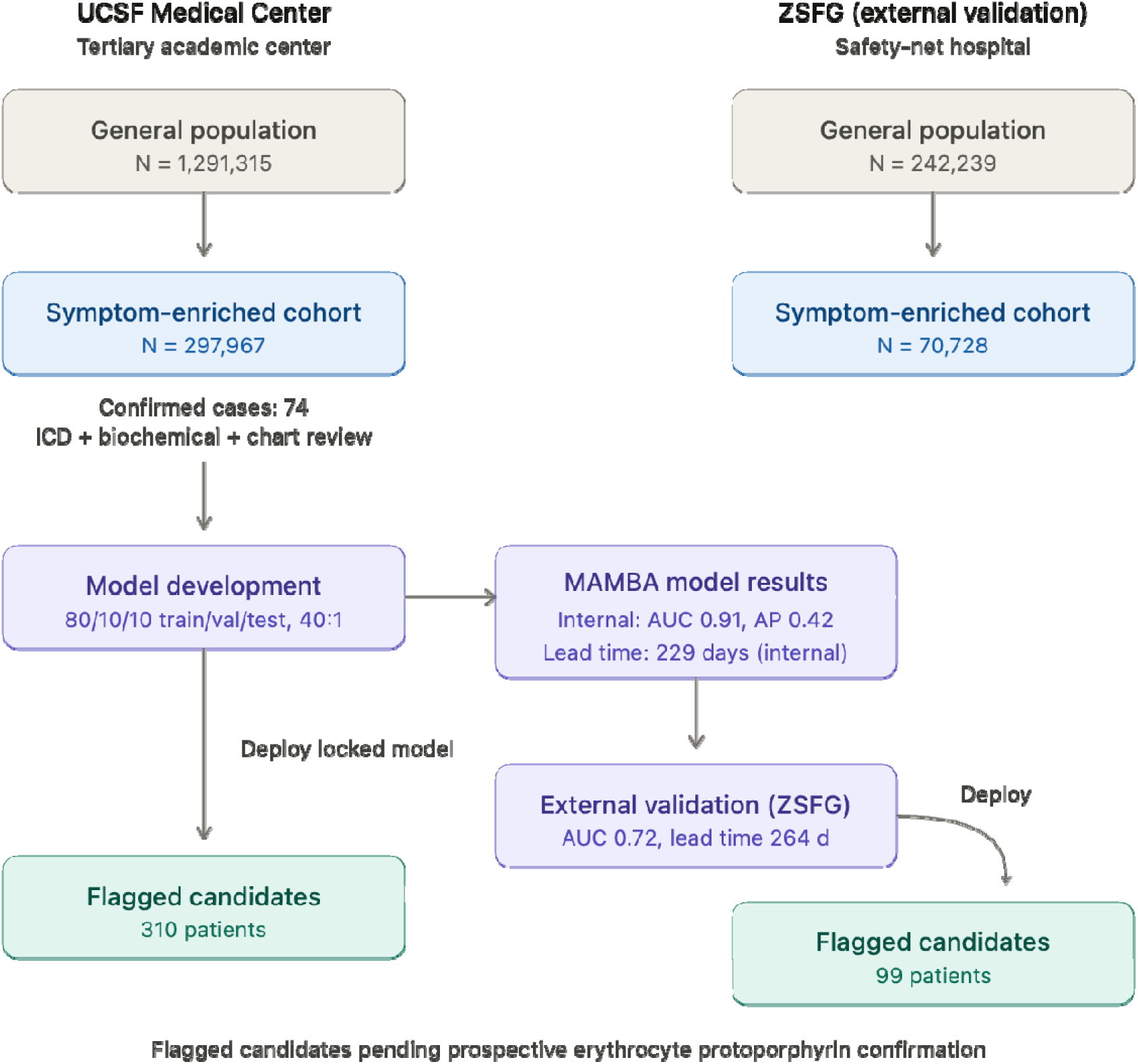
Study flow diagram. Schematic of case ascertainment, symptom-enriched control selection, feature extraction, model development, and deployment cascades at UCSF and ZSFG. At UCSF, the general patient population (N = 1,291,315) is progressively filtered to patients with EPP-compatible symptoms (N = 297,967), then to model-flagged high-risk patients (n = 310), and finally to an estimated ∼130 expected true positives. The parallel cascade at ZSFG begins with 242,239 patients and yields 99 flagged patients and ∼10 expected true positives.

**Figure 2:**
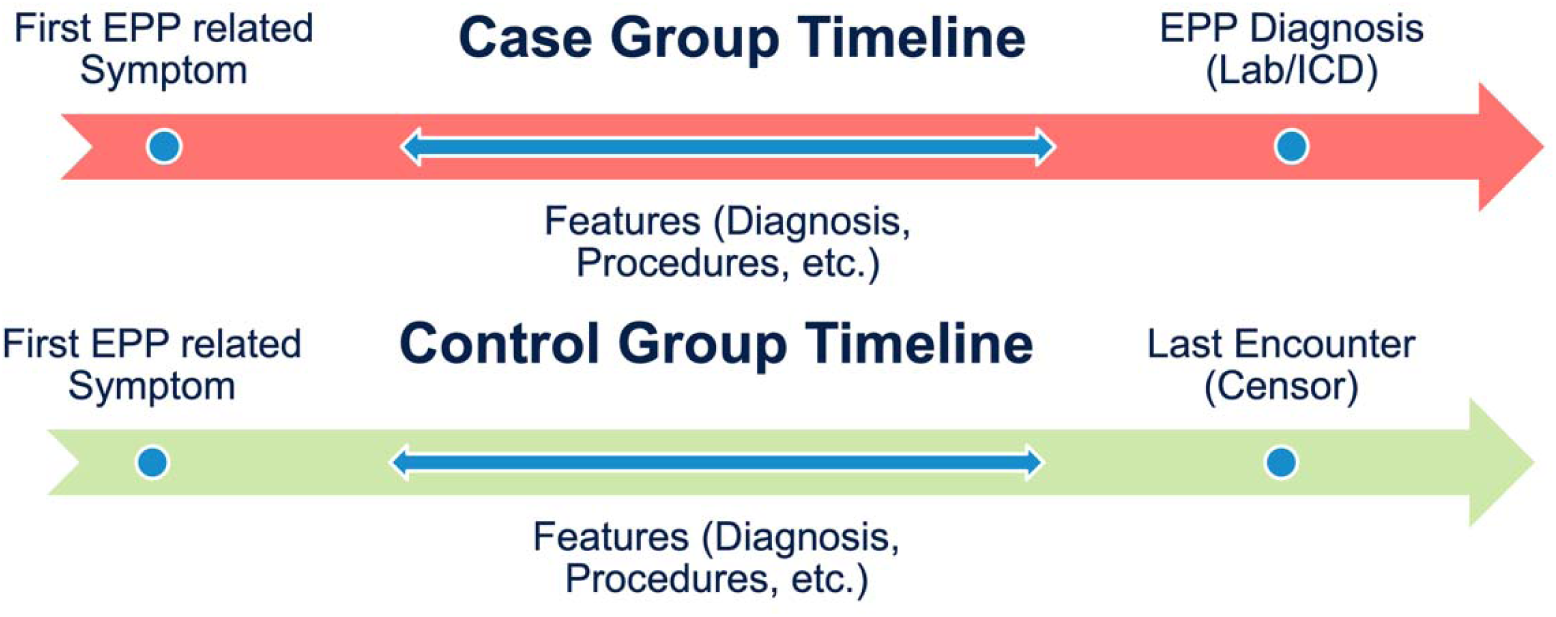
Timeline illustrating feature extraction for cases and controls. For EPP cases, clinical features were collected from the first EPP-related symptom to the diagnosis date. For controls, features were collected from the first EPP-related symptom to the last EPP free clinical encounter or death.

### 2.9 External Validation

External validation was performed on the ZSFG patient population, using the locked UCSF-trained model without any retraining or threshold adjustment. The ZSFG data warehouse was queried using an identical feature extraction pipeline. Performance metrics were computed within a ZSFG test cohort of symptom-enriched patients, and the deployed model was then applied to the full ZSFG symptom-compatible population (N = 70,728) to estimate the number of potentially undiagnosed individuals (Figure 1).

### 2.10 Statistical Analysis

Descriptive statistics were reported as counts and percentages for categorical variables and as means with standard deviations for continuous variables. Confidence intervals were not computed for model metrics given the single fixed test set; performance was instead reported alongside cross-validated training estimates. Analyses were conducted in Python using scikit-learn, CatBoost, PyTorch, and the MAMBA reference implementation.

## 3. Results

### 3.1 Cohort Characteristics

At UCSF, 74 confirmed EPP cases were identified through the combined ICD-based and biochemical/clinical review strategy described above. The mean age at diagnosis was 33.7 years (SD 20.8), 57.7% of cases were female, and the case population was ethnically diverse, with 57.7% identifying as White, 12.7% as Latinx, 4.2% as Asian, and 2.8% as Southwest Asian and North African (Table 1). The held-out test set used for model evaluation comprised 1,865 patients, of whom 43 were confirmed EPP cases and 1,822 were symptom-enriched controls, consistent with the prespecified 40:1 control-to-case ratio. Eligible controls carried at least one of the symptom-based diagnosis codes shown in Table 1, spanning abdominal, dermatologic, hematologic, hepatic, and ocular presentations.

**Table 1:** Most common EPP-related symptom ICD codes in the study population.

|  |  |  |
| --- | --- | --- |
| ICD group | ICD description | n (%) |
| R10 | Abdominal and pelvic pain | 360,182 (10.7 %) |
| D64 | Anemia, unspecified and other anemias | 206,739 (6.1 %) |
| R21 | Rash and other nonspecific skin eruption | 167,517 (5.0 %) |
| H53 | Visual disturbances | 98,997 (2.9 %) |
| K76 | Other diseases of liver (e.g., fatty liver, hepatic fibrosis, cirrhosis) | 87,619 (2.6 %) |
| L98 | Other disorders of skin and subcutaneous tissue | 84,155 (2.5 %) |
| D50 | Iron deficiency anemia | 74,576 (2.2 %) |
| L29 | Pruritus (itching) | 73,860 (2.2 %) |
| L57 | Skin changes due to chronic exposure to nonionizing radiation (e.g., actinic keratosis) | 55,821 (1.7 %) |
| L90 | Atrophic disorders of skin (e.g., scar, striae) | 45,599 (1.4 %) |
| L85 | Disorders of keratinization (e.g., lichenification, keratosis) | 44,607 (1.3 %) |
| L50 | Urticaria (hives) | 44,510 (1.3 %) |
| K80 | Cholelithiasis (gallstones) | 37,024 (1.1 %) |
| L91 | Hypertrophic disorders of skin (e.g., keloid, hypertrophic scar) | 26,819 (0.8 %) |
| K82 | Other diseases of gallbladder | 14,196 (0.4 %) |
| K81 | Cholecystitis (acute or chronic) | 8,982 (0.3 %) |
| L56 | Other acute skin changes due to UV radiation (e.g., sunburn) | 4,700 (0.1 %) |
| K77 | Liver disorders in diseases classified elsewhere | 197 (0.006 %) |

### 3.2 Internal Model Performance

On the held-out UCSF test set, the CatBoost model achieved an AUC–ROC of 0.89 and an average precision of 0.27 (Figure 3). Overall accuracy was 93.3%, sensitivity 62.8%, specificity 94.0%, precision 19.9%, and F1 score 0.30. The MAMBA sequence model demonstrated higher discrimination and substantially higher precision: AUC–ROC 0.91, average precision 0.42, accuracy 98.2%, sensitivity 46.5%, specificity 99.4%, precision 64.5%, and F1 score 0.54 (Table 2). The substantial improvement in precision–recall performance from CatBoost to MAMBA indicates that the sequence model more effectively separates true cases from symptom-matched controls, which is the clinically relevant axis for a rare disease screening tool.

**Figure 3.**
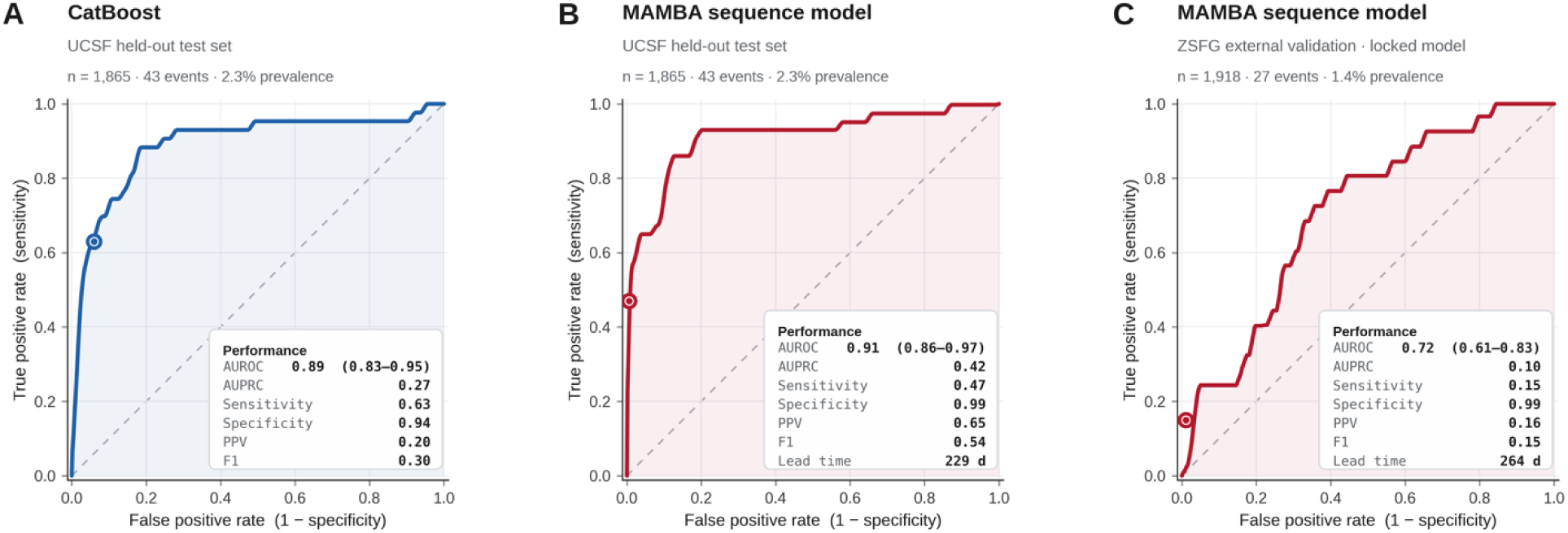
Receiver operating characteristic curves and performance metrics. (A) CatBoost model on the UCSF held-out test set (n = 1,865): AUC–ROC 0.89, average precision 0.27. (B) MAMBA sequence model on the same UCSF test set: AUC–ROC 0.91, average precision 0.42, with a median diagnostic lead time of 229 days. (C) External validation of the locked MAMBA model at ZSFG without retraining: AUC–ROC 0.72, average precision 0.10, with a median diagnostic lead time of 264 days. Diagnostic lead time is the median interval between the model’s first high-risk prediction and the documented clinical diagnosis among true-positive cases.

**Table 2.** Model performance on the UCSF held-out test set (n = 1,865; 43 cases, 1,822 controls)

| Metric | CatBoost | MAMBA |
| --- | --- | --- |
| Accuracy | 0.933 | 0.982 |
| Sensitivity (recall) | 0.628 | 0.465 |
| Specificity | 0.940 | 0.994 |
| Precision (PPV) | 0.199 | 0.645 |
| F1 score | 0.302 | 0.541 |
| AUC–ROC | 0.890 | 0.913 |
| Average precision | 0.270 | 0.424 |
| Median diagnostic lead time (days) | — | 229 |

*AUC–ROC, area under the receiver operating characteristic curve; PPV, positive predictive value. Diagnostic lead time was computed for the best-performing model only.*

### 3.3 Diagnostic Lead Time

Among true-positive cases in the UCSF test set, the MAMBA model’s first high-risk classification preceded documented EPP diagnosis by a median of 7.5 months. This indicates that for most cases detected by the model, a clinically meaningful opportunity window exists during which confirmatory erythrocyte protoporphyrin testing could have been initiated prior to formal diagnosis.

### 3.4 Population-Level Deployment at UCSF

Deployment of the locked MAMBA model across the UCSF population with any encounter in the preceding three years and at least one EPP-compatible symptom code (N = 297,967) produced 310 high-risk predictions. These 310 flagged patients represent a clinically tractable list for prospective confirmatory testing.

The demographic profile of flagged patients differed modestly from that of the known UCSF EPP case population: 53% were male, 39% identified as Latinx, 17% as Black or African American, 12% as White, and 10% as Asian, with a distribution skewed toward younger age groups (the 0–4 year category accounting for the largest bin; Table 4). Among flagged patients, the most frequent symptom codes were other anemias (70.0%), iron deficiency anemia (49.7%), and abdominal and pelvic pain (41.0%).

### 3.5 External Validation at ZSFG

External validation was performed on the ZSFG patient population using the locked UCSF-trained MAMBA model. In the ZSFG test cohort, performance attenuated relative to internal validation: AUC–ROC 0.72 (Figure 3), average precision 0.098, accuracy 97.7%, sensitivity 16.0%, specificity 98.8%, and precision 14.8% (Table 3). Among the true EPP cases identified by the model before their actual diagnosis date, the model flagged them a median of 264 days earlier.

**Table 3.** External validation of the UCSF-trained MAMBA model at ZSFG.

| Metric | Value |
| --- | --- |
| Patients correctly labeled | 1,874 |
| Patients incorrectly labeled | 44 |
| Accuracy | 0.977 |
| Sensitivity (recall) | 0.160 |
| Specificity | 0.988 |
| Precision (PPV) | 0.148 |
| AUC–ROC | 0.717 |
| Average precision | 0.098 |
| False positive rate | 0.012 |
| Median diagnostic lead time (days) | 264 |

*Confusion matrix at the locked operating threshold: true positives 4; false positives 23; true negatives 1,870; false negatives 21.*

Applied to the broader ZSFG symptom-compatible population (N = 70,728 of 242,239 patients with any encounter), the model flagged 99 additional patients as high risk (> 50% probability).

The flagged ZSFG population differed notably in demographic composition, demonstrating some model generalizability: 56% identified as Hispanic/Latino, 27% as Black or African American, 8% as White, and 5% as Asian, and 78% were female (Table 4). The age distribution was skewed toward young and middle adulthood rather than early childhood, with peaks in the 20–34 year range. The dominant symptom codes among flagged ZSFG patients were iron deficiency anemia (82/99), abdominal and pelvic pain (57/99), and other anemias (53/99).

**Table 4.** Demographic and clinical characteristics of model-flagged high-risk patients at UCSF and ZSFG.

| Characteristic | UCSF (n = 310) | ZSFG (n = 99) |
| --- | --- | --- |
| Female, n (%) | 146 (47) | 77 (78) |
| Male, n (%) | 164 (53) | 21 (21) |
| Hispanic/Latinx, n (%) | 122 (39) | 55 (56) |
| Black/African American, n (%) | 52 (17) | 26 (27) |
| White, n (%) | 37 (12) | 8 (8) |
| Asian, n (%) | 32 (10) | 5 (5) |
| Iron deficiency anemia, n (%) | 154 (49.7) | 82 (83) |
| Other anemias, n (%) | 217 (70.0) | 53 (54) |
| Abdominal/pelvic pain, n (%) | 127 (41.0) | 57 (58) |
| Rash or nonspecific skin eruption, n (%) | 85 (27.4) | 26 (26) |
| Ever had protoporphyrin lab, n (%) | 44 (14.2) | 14 (14) |

*Row totals may not sum to population size because patients may identify as multiple race/ethnicities or carry multiple symptom-compatible diagnoses. Columns reflect multi-label counts.*

## 4. Discussion

In this two-site retrospective case–control study, we developed and externally validated machine learning models for early identification of EPP from longitudinal EHR data. The sequence-based MAMBA model outperformed a gradient boosting baseline on the internal test set, achieving an AUC–ROC of 0.91 and an average precision of 0.42 in a deliberately complex, symptom-enriched control population. Among true cases, the model identified high-risk status a median of 229 days before documented diagnosis, and when deployed health–system–wide, it flagged 310 patients, implying a clinical prevalence closer to published genetic estimates than to historical clinical estimates. External validation in a demographically and socioeconomically distinct safety-net hospital demonstrated meaningful, if attenuated, performance (AUC–ROC 0.72), a preserved ability to flag cases well in advance of formal diagnosis, and similar clinical prevalence.

### 4.1 Clinical Importance of Earlier Detection

EPP is characterized by severe, non-blistering photosensitivity that typically begins in early childhood, together with variable risk of progressive hepatobiliary disease.^2–4^ Because cutaneous symptoms often lack visible skin findings and because systemic manifestations are nonspecific, patients commonly experience multi-year diagnostic delays during which they carry alternative labels of anemia, abdominal pain, functional dermatoses, or unexplained liver abnormalities. Earlier recognition enables targeted biochemical confirmation with erythrocyte protoporphyrin testing, initiation of photoprotection counseling, consideration of afamelanotide therapy where appropriate,^14^, and systematic hepatic surveillance.

Our findings indicate that the phenotypic signature preceding EPP diagnosis is detectable in routine structured EHR data months before clinicians converge on the correct diagnosis. The dominant features in flagged patients, such as iron-related anemias, abdominal pain, nonspecific rash, and hepatobiliary abnormalities, track closely with clinical descriptions of pre-diagnosis EPP. And our sequence-based model demonstrated the ability to integrate the temporal co-occurrence of these features, outperforming a static feature-based approach.

### 4.2 Underdiagnosis and Epidemiologic Implications

A central observation of this work is the magnitude of implied underdiagnosis. If the deployed UCSF model’s positive predictive value (∼0.42) holds among the 310 high-risk patients it flagged, the corresponding implied prevalence is roughly an order of magnitude higher than traditional clinical prevalence estimates and broadly consistent with the UK Biobank genetic prevalence estimate of ∼1 in 17,000.^7^ A similar pattern is seen at ZSFG, where the implied prevalence of approximately 1 in 24,000 also materially exceeds historical clinical figures. Taken together, these estimates are consistent with the hypothesis that EPP is systematically underdiagnosed in real-world clinical practice and that a large fraction of genetically susceptible individuals remain undetected, even at academic centers with expertise in porphyria.

### 4.3 Comparison with Prior Work

Most prior EPP literature has focused on pathophysiology, genetic characterization, clinical management, and natural history rather than systematic case finding.^1–6^ Population-based genetic work, notably the UK Biobank study by Dickey and colleagues, first quantified the gap between genetic and clinical prevalence for EPP.^7^ Our study complements this line of work by showing that a substantial fraction of this gap is addressable from within routine clinical data alone, without recourse to genotyping. More broadly, prior EHR-based rare disease detection efforts for acute hepatic porphyrias, primary immunodeficiencies, and inborn errors of metabolism have converged on a common finding: rare diseases leave detectable longitudinal phenotypic signatures in structured data.^8–11^ Our work extends this paradigm specifically to EPP and, importantly, includes external validation across a socioeconomically distinct hospital system rather than only a train/test split from the same population.

### 4.4 Generalization Across Health Systems

The attenuation of model performance at ZSFG merits careful interpretation. Several factors plausibly contribute: (a) the UCSF case-defining population is referred, while ZSFG serves a primarily safety-net, publicly insured, and underserved patient population; (b) documentation density, laboratory ordering patterns, and specialty access differ substantially between the two settings; and (c) the dominant clinical phenotype of flagged ZSFG patients is shifted toward adulthood and iron deficiency anemia rather than early childhood photosensitivity, suggesting the model may be capturing late-presenting rather than archetypal EPP in this population. This pattern is consistent with a growing literature on the generalizability of clinical AI and reinforces the need for local recalibration, threshold adjustment, and site-specific validation prior to operational deployment.^15–16^

### 4.5 Clinical and Public Health Implications

The implications of this work extend beyond EPP. Rare liver-associated and metabolic disorders frequently escape recognition because their individual presentations overlap with common conditions. A scalable EHR-based screening approach, applied at the health-system or network level, could flag a small, clinically manageable number of high-risk patients for targeted biochemical testing—an intervention that is low-cost, minimally invasive, and directly actionable. In hepatology and rare disease care specifically, such an approach may shift the operational posture from reactive diagnosis at the point of severe complications to proactive case finding months to years earlier. Improved case ascertainment would also refine prevalence estimates and support epidemiologic surveillance of conditions for which disease-modifying therapies are now available or under investigation.

### 4.6 Strengths

This study has several strengths. First, case ascertainment combined administrative coding with biochemical confirmation and specialty chart review, reducing reliance on any single, fallible source. Second, the symptom-enriched control design reflects the operational population in which such a model would actually need to discriminate, rather than an artificially easy comparator. Third, evaluation included not only internal test-set performance but also population-level deployment and external validation in a demographically distinct site which is an uncommon combination in the rare disease modeling literature.

### 4.7 Limitations

Several limitations warrant emphasis. First, the retrospective design depends on the accuracy and completeness of structured EHR data; misclassification of either cases or controls is possible despite multiple ascertainment strategies. Second, although precision–recall performance was strong internally, sensitivity was modest, reflecting the well-known trade-off in rare disease detection at low prevalence. Third, the externally validated operating point yielded limited positive predictive value at ZSFG, so any operational deployment in such settings would require threshold recalibration and, ideally, local fine-tuning. Fourth, the flagged individuals have not undergone prospective confirmatory testing; estimates of true-positive yield therefore remain model-derived rather than laboratory-confirmed. Finally, this work did not evaluate cost-effectiveness or clinical outcomes associated with an earlier diagnosis, questions that require prospective implementation studies.

### 4.8 Future Directions

The logical next step is prospective evaluation. A pragmatic pilot in which flagged patients receive clinician-reviewed outreach for confirmatory erythrocyte protoporphyrin testing would directly estimate true-positive yield and clinical impact. Methodologically, incorporating unstructured notes, especially using language models, could improve sensitivity for poorly coded photosensitivity symptoms. Local recalibration on safety-net EHR data is likely essential for equitable deployment. More broadly, this pipeline can be extended to other rare hepatologic and metabolic disorders that are challenged by a simmilar prolonged diagnostic delay.

## 5. Conclusion

Longitudinal EHR-based machine learning can identify patients with erythropoietic protoporphyria months before formal clinical diagnosis, revealing a detectable phenotypic signature of an underrecognized disease and corroborating genetic evidence of substantial underdiagnosis in routine care. The sequence-based MAMBA model substantially outperformed a gradient boosting baseline internally, generalized with expected attenuation, to a demographically distinct safety-net hospital, and flagged hundreds of previously undetected high-risk individuals at implied prevalences approaching genetic estimates. Continued refinement, prospective validation, and equitable, site-adapted deployment of such tools have the potential to shorten diagnostic delay and advance precision approaches to rare disease care in hepatology.

## Acknowledgments

The authors acknowledge Hunter Mills MS, Habibeh Ashouri Choshali PhD, and Yuvraaj Kapoor BS for their contributions to prior versions of this work.

## Data availability

Deidentified electronic health records data from UCSF and ZSFG are available to support study reproducibility and extensions following the execution of a data use agreement with these health systems.

## Contributions

A.A.: conceptualization, data curation, formal analysis, investigation, methodology, software, validation, visualization, writing – original draft, and writing – review & editing. G.O.: formal analysis, investigation, methodology, software, and visualization. A.S.: data curation, investigation, resources, and software. B.W.: conceptualization, funding acquisition, investigation, methodology, supervision, validation, and writing – review & editing. V.A.R.: conceptualization, formal analysis, funding acquisition, investigation, supervision, validation, and writing – review & editing. S.A.: data curation, resources, and validation. All authors read and approved the final manuscript.

## Ethics Approval

This study was approved by the University of California, San Francisco (UCSF) Institutional Review Board (IRB #24-42109), which granted a waiver of informed consent and HIPAA authorization for retrospective chart review. The research was determined to pose no greater than minimal risk to participants.

## References

1. Balwani M, Naik H, Anderson KE, et al. Clinical, biochemical, and genetic characterization of North American patients with erythropoietic protoporphyria and X-linked protoporphyria. JAMA Dermatol. 2017;153(8):789–796.

2. Lecha M, Puy H, Deybach JC. Erythropoietic protoporphyria. Orphanet J Rare Dis. 2009;4:19.

3. Balwani M. Erythropoietic protoporphyria and X-linked protoporphyria: pathophysiology, genetics, clinical manifestations, and management. Mol Genet Metab. 2019;128(3):298–303.

4. Anderson KE, Bloomer JR, Bonkovsky HL, et al. Recommendations for the diagnosis and treatment of the acute porphyrias. Ann Intern Med. 2005;142(6):439–450.

5. Holme SA, Anstey AV, Finlay AY, Elder GH, Badminton MN. Erythropoietic protoporphyria in the U.K.: clinical features and effect on quality of life. Br J Dermatol. 2006;155(3):574–581.

6. Wensink D, Wagenmakers MAEM, Langendonk JG. Afamelanotide for prevention of phototoxicity in erythropoietic protoporphyria. Expert Rev Clin Pharmacol. 2021;14(2):151–160.

7. Dickey AK, Quick C, Ducamp S, et al. Evidence in the UK Biobank for the underdiagnosis of erythropoietic protoporphyria. Genet Med. 2021;23(1):140–148.

8. Cohen AM, Chamberlin S, Deloughery T, et al. Detecting rare diseases in electronic health records using machine learning and knowledge engineering: case study of acute hepatic porphyria. PLoS One. 2020;15(7):e0235574.

9. Colbaugh R, Glass K, Rudolf C, Volk MT. Learning to identify rare disease patients from electronic health records. AMIA Annu Symp Proc. 2018;2018:340–347.

10. Rafee A, Riepenhausen S, Neuhaus P, et al. Rare2Common: A reasoning framework for rare disease decision support using electronic health records. J Biomed Inform. 2022;128:104035.

11. Shen F, Liu S, Wang Y, Wen A, Wang L, Liu H. Utilization of electronic medical records and biomedical literature to support the diagnosis of rare diseases using data fusion and collaborative filtering approaches. JMIR Med Inform. 2018;6(4):e11301.

12. Prokhorenkova L, Gusev G, Vorobev A, Dorogush AV, Gulin A. CatBoost: unbiased boosting with categorical features. Adv Neural Inf Process Syst. 2018;31:6638–6648.

13. Gu A, Dao T. Mamba: Linear-time sequence modeling with selective state spaces. arXiv:2312.00752. 2023.

14. Langendonk JG, Balwani M, Anderson KE, et al. Afamelanotide for erythropoietic protoporphyria. N Engl J Med. 2015;373(1):48–59.

15. Finlayson SG, Subbaswamy A, Singh K, et al. The clinician and dataset shift in artificial intelligence. N Engl J Med. 2021;385(3):283–286.

16. Wong A, Otles E, Donnelly JP, et al. External validation of a widely implemented proprietary sepsis prediction model in hospitalized patients. JAMA Intern Med. 2021;181(8):1065–1070.

17. Collins GS, Reitsma JB, Altman DG, Moons KGM. Transparent Reporting of a Multivariable Prediction Model for Individual Prognosis or Diagnosis (TRIPOD): The TRIPOD Statement. Ann Intern Med. 2015;162(1):55–63.

